# DIFFERENTIAL SARS-COV-2 ANTIGEN SPECIFICITY OF THE HUMORAL RESPONSE IN INACTIVATED VIRUS-VACCINATED, CONVALESCENT, AND BREAKTHROUGH SUBJECTS

**DOI:** 10.1101/2022.07.01.22277165

**Authors:** Luisa F. Duarte, Yaneisi Vázquez, Benjamín Diethelm-Varela, Valentina Pavez, Roslye Berríos-Rojas, Jessica A. White, Alexis M. Kalergis, Susan M. Bueno, Pablo A. González

## Abstract

Analytical methods for the differential determination between natural infection with SARS-CoV-2 vs. immunity elicited by vaccination or infection after immunization (breakthrough cases) represent attractive new research venues in the context of the ongoing COVID-19 pandemic caused by Severe Acute Respiratory Syndrome coronavirus 2 (SARS-CoV-2). Herein, we set out to compare humoral responses against several SARS-CoV-2 structural and non-structural proteins in infected unvaccinated (convalescent), vaccinated, as well as vaccinated and infected (breakthrough) individuals. Our results indicate that immunization with an inactivated SARS-CoV-2 vaccine (CoronaVac) induces significantly higher levels of IgG antibodies against the membrane (M) protein of SARS-CoV-2 as compared to convalescent subjects both, after the primary vaccination schedule and after a booster dose. Moreover, we found that CoronaVac-immunized individuals, after receiving a third vaccine shot, display equivalent levels of N-specific IgG antibodies as convalescents subjects. Regarding non-structural viral proteins, for the two viral proteins ORF3a and NSP8, IgG antibodies were produced in more than 50% of the convalescent subjects. Finally, a logistic regression model and a receiver operating characteristic (ROC) analysis show that combined detection of M and N proteins may be useful as a biomarker to differentiate breakthrough cases from vaccinated and convalescent individuals that did not receive prior vaccination. Taken together, these results suggest that multiple SARS-CoV-2 antigens may be used as differential biomarkers for distinguishing natural infection from vaccination.

## 1. INTRODUCTION

Severe Acute Respiratory Syndrome coronavirus 2 (SARS-CoV-2), the etiological agent of coronavirus disease 2019 (COVID-19), produces significant morbidity and mortality worldwide that has caused significant strain on public health systems, altogether imposing severe social and economic burdens to nations around the globe [1, 2]. Starting the year 2021, most countries worldwide have deployed massive vaccination campaigns that successfully curb the worst outcomes of COVID-19. However, the immune responses elicited in convalescent individuals, the vaccinated, and those vaccinated and infected (breakthrough) cases remain to be studied in depth in the upcoming years in such a way to identify and better understand potential correlates of protection against infection with this virus [3]. Worldwide, more than a dozen vaccines have been approved by the World Health Organization (WHO), which are based on different technological platforms, compositions and administration schedules [4, 5]. Importantly, whole inactivated SARS-CoV-2 vaccines account for nearly half of the total number of doses administrated in the world population, and CoronaVac (Sinovac Biotech) has been approved in over 30 countries within mass vaccination campaigns in low and middle-income countries [6], with favorable safety and immunogenicity results [7–10]. The efficacy and effectiveness of this vaccine against COVID-19-related hospitalizations and deaths are significant [11–16], especially when immunity is reinforced with a booster dose [17–20].

Whole-virus vaccines have a variety of structural viral antigens, many of which are not present in mRNA- or vector-based vaccines [7]. Importantly, whole-virus vaccines may elicit, to some extent, immune responses that mimic natural infection with SARS-CoV-2. However, because the profile of antigenic proteins presented in the context of infectious virus and contained in the inactivated viral particles within the vaccine are not identical, we hypothesized that natural infection versus inactivated-virus vaccination or breakthrough cases elicits differential humoral responses against SARS-CoV-2 proteins. Thus, we sought to identify these putative differences, which could eventually be used as biomarkers for the differential serodiagnosis of individuals naturally infected with SARS-CoV-2 versus those immunized with inactivated-virus vaccine or breakthrough cases undergoing infection after vaccination.

The SARS-CoV-2 genome encodes four structural proteins, namely Spike (S), nucleocapsid (N), membrane (M), and envelope (E), and 16 non-structural proteins (NSP1- NSP16), along with several accessory proteins (ORF3a, 3d, 6, 7a, 7b, 8, 9b, 14, and 10) that play key roles in immune evasion and the replication cycle of the virus [21, 22]. Among these proteins, the viral structural protein S is highly immunogenic, and the induction of binding- and neutralizing anti-S antibodies by vaccines against SARS-CoV-2 have been the focus of extensive investigation during the last two years [23–27]. However, antibodies against other SARS-CoV-2 proteins, such as those produced against virus-inactivated vaccines, are less known and thus, require further characterization for a better understanding of the kinetics and extent of the host immune profiles generated against this virus and potentially identifying correlates of protection.

In this study, we compared the humoral immune responses elicited against the N, M, and E-structural proteins of SARS-CoV-2, as well as the non-structural ORF3a, and NSP8 proteins of this virus among unvaccinated convalescent individuals that survived SARS-CoV-2 infection, and subjects immunized with the SARS-CoV-2-inactivated CoronaVac vaccine at different stages during their vaccination and booster schedule. We also analyzed and compared this humoral response in CoronaVac-vaccinated individuals later infected with SARS-CoV-2 (breakthrough cases) [17]. Overall, our data show that anti-N IgG antibodies are produced significantly in the vaccinated group only after administering a booster dose of CoronaVac, and anti-M IgG antibody titers were higher in the vaccinated group as compared to convalescent individuals. Conversely, convalescent individuals displayed high titers of anti-N antibodies, which remained elevated for at least eight weeks after recovery from SARS-CoV-2 infection. These individuals also displayed a seropositivity rate above 50% for the NSP8 protein. Regarding breakthrough cases, an antibody pattern characteristic of an anamnestic immune response was elicited, resulting in a substantial boost of titers against surface antigens within the virus. Finally, we performed a receiver operating characteristic (ROC) analysis to differentiate breakthrough cases between vaccinated volunteers and convalescent individuals without previous vaccination. Importantly, we found at the protein level that the antibody responses against the combination of the N and M viral proteins may serve to discriminate between these individuals with an appropriate cutoff criterion.

## 2. MATERIALS AND METHODS

### **2.1** Subjects

Serum samples from subjects between 18 to 59 years and older than 60 years belonging to the CoronaVac03CL Phase 3 scientific-clinic study carried out in Chile were obtained from the following groups: i) pre-immune: sera obtained at the moment of the first dose, ii) second dose + 2 weeks: sera obtained from subjects vaccinated with the second dose of CoronaVac (28 days post-first dose) plus two weeks, iii) second dose + 4w: sera from subjects vaccinated with the second dose of CoronaVac plus four weeks, iv) third dose + 4w: sera from subjects vaccinated with the booster dose of CoronaVac (6 months after the first dose) plus four weeks, vi) breakthrough + 2w: serum obtained from subjects vaccinated with two doses of CoronaVac and then naturally infected with SARS-CoV-2, from whom the samples were taken two weeks after a positive polymerase chain reaction (PCR) test, and vii) breakthrough + 4w: serum from subjects vaccinated with two doses of CoronaVac and infected with SARS-CoV-2, from whom the sample was taken four weeks after a positive PCR test. At least 25 subjects were evaluated for the vaccinated group and 10 for the breakthrough cases. Sera obtained from the biobank of clinical samples within the PATH institution was grouped as follows: i) naive: sera from 10 subjects who were not exposed to the virus and were taken before the year 2020, ii) convalescents: sera from 9 subjects who were infected with SARS-CoV-2 and then blood samples were taken for serum recollection at 1, 2, 4 and 8 weeks after symptom-onset, iii) 10 samples obtained from convalescent subjects that were classified with low, mid and high titer against SARS-CoV-2, but the time of the sample collection was not detailed (unspecified). The age of these subjects was unknown. Table 1 summarizes the description of all samples included in this study. Blood samples were obtained from volunteers recruited in the clinical trial CoronaVac03CL (clinicaltrials.gov #NCT04651790) carried out in Chile starting in November 2020. The study was approved by the sponsoring institution Ethical Committee (ID 200708006), each Institutional Ethical Committee of the other sites, and the Public Health Institute of Chile (ISP Chile, number 24204/20). Execution of the trials was conducted according to the current Tripartite Guidelines for Good Clinical Practices, the Declaration of Helsinki and local regulations.

**Table 1.**

| Group | Time point evaluated | Gender | Age | Total |
| --- | --- | --- | --- | --- |
| **Vaccinated** | Pre-immune<br>2nd dose + 2 weeks<br>2nd dose + 4 weeks<br>3rd dose + 4 weeks | Female | 18-59 years | 12 |
|  |  |  | > 60 years | 4 |
|  |  | Male | 18-59 years | 5 |
|  |  |  | > 60 years | 5 |
| **Convalescents** | 1 week<br>2 weeks<br>4 weeks<br>8 weeks | Unknown | Unknown | 10 |
|  | Unspecified | Unknown | Unknown | 9 |
| **Breakthrough** | 2 weeks<br>4 weeks | Female | 18-59 years | 3 |
|  |  |  | > 60 years | 0 |
|  |  | Male | 18-59 years | 4 |
|  |  |  | > 60 years | 3 |
| **Naïve** | Before the year 2020 | Unknown | Unknown | 10 |

### **2.2** Dot blot

To immobilize recombinant proteins on a solid matrix, 500 ng of each protein was diluted in 20 mM phosphate buffer (mixture of monobasic and dibasic phosphate), 0.5 M NaCl, and 8 M urea were spotted onto a nitrocellulose membrane (in 2 uL volume) (Thermo Scientific). Membranes with spots were air-dried for 15 min and subsequently blocked with 10% BSA diluted in 0.05% Tween-20 in PBS, for 2 h at RT. After incubation, the membranes were washed with 0.05% Tween-20 in PBS (twice). Next, the membranes were incubated overnight at 4°C with a sera pool from naive, convalescents (+ 4 weeks), vaccinated with second dose + 2 weeks, and breakthrough (PCR(+) + 2 weeks) subjects. A sera pool was incubated in a dilution of 1/250 with 1% BSA diluted in 0.05% Tween-20 in PBS. As positive control, all proteins (500 ng) were incubated with an anti-His Tag antibody conjugated with biotin at a dilution of 1:3,000 in 1% BSA diluted in 0.05% Tween-20 in PBS (Supplementary Figure 1). After incubation, the membranes were washed with 0.05% Tween-20 in PBS (three times x 5 min) and incubated for 1 h at RT with 1:2,000 anti-human IgG-HRP (1 mg/ml, BD) diluted in 1% BSA with 0.05% and Tween-20 in PBS. The membranes with an anti-His Tag- biotin antibody (Genscript, # A00613) were incubated with Streptavidin-HRP (1:6,000; Abcam #7403). Finally, membranes were washed with 0.05% Tween-20 in PBS (three times), once with PBS, and then incubated with an enhanced chemiluminescence Western blot detection system (Femto, ECL, Thermo Scientific # 34094). Proteins evaluated included ORF1a polyprotein (Invitrogen, #RP-87701), ORF3a (LSBio, #LS-G145920), ORF8 (LSBio, #LS-G145919), NSP1 (R&D systems, #10666-CV), NSP8 (R&D systems, #10633- CV), NSP9 (R&D systems, #10631-CV), NSP10 (R&D systems, #10630-CV), NSP14 (R&D systems, #10667-CV), E (Sino Biological, #40609-V10E3) and ME (Sino Biological, #40598-V07E).

### **2.3** ELISA assays

In-house indirect ELISA assays were performed to detect humoral immunity against N-SARS-CoV-2 (R&D systems, #10474-CV), M (R&D systems, #10690-CV), E (Sino Biological, #40609-V10E3), ORF3a (LSBio, #LS-G145920) and NSP8 (R&D systems, #10633-CV). Briefly, high-binding 96-well ELISA plates (Corning, #9018) were activated with 100 ng of N and M antigen dissolved in carbonate-bicarbonate buffer (Bioleged 1x, #421701) for 1 h at 37°C and blocked with 10% m/w milk in PBS 1X - Tween 20 (0.05%) overnight at 4°C. Denaturing buffer (20 mM monobasic and dibasic phosphate buffer, 0.5 NaCl, and 8 M urea) was used as co-acting buffer to evaluate anti-ORF3a, anti-E, and anti- NSP8-SARS-CoV-2 (250 ng, 500 ng and 200 ng, respectively). Afterward, the plates were incubated with sera from participants using serial dilution factors ranging from 1/250 to 1/16,000 (N and M) and 1/50 to 1/1,600 (E, NSP8, and ORF-3a proteins) for 1 h at 37°C (diluted 1% m/w milk in PBS 1X - Tween 20 (0.05%)). In parallel, a WHO standard curve (NIBSC code: 20/268) was performed from dilutions of 1/40 to 1/360. Then, the plates were incubated with anti-human IgG-HRP (BD, # 555788) for 30 min at room temperature in darkness. Finally, plates were resolved using commercial TMB (BD OptEIA, # 555214), consisting of a substrate mixture, which was incubated at room temperature for 15 min in darkness. This reaction was stopped using 2N H2SO4, and the absorbance at 450 nm was read.

### **2.4** Data analysis

For all ELISA assays, corrected optical density (OD) values were calculated by obtaining the average between each subject’s two replicates and subtracting the corresponding subject’s blank (i.e., the OD measurement from the “inactivated” well) for each dilution factor. An absorbance cutoff was calculated for each dilution factor to establish the seropositivity threshold. For each subject, the antibody titer was defined as the highest dilution factor where the corrected OD was higher than the cutoff value for the corresponding dilution factor. Geometric Mean of Titers (GMTs) was calculated using all subject titers for a given visit. A standard curve was used to plot the antibody responses against the M, N, and NSP8 proteins in the samples as binding arbitrary units (BAU), by using the WHO International Standard for SARS-CoV-2 antibody (NIBSC code: 20/268), which was prepared according to the manufacturer’s instructions [28]. Data were analyzed using the log concentration transformed, and the final concentration for each sample was the average of the product of the interpolated BAU from the standard curve and the sample dilution factor required to achieve the OD450 value that fell within the linear quantitative range. Samples with undetermined concentrations at the lowest dilution tested were assigned the lower limit of quantification (28.6; 23.6; and 13.5 BAU for N, M, and NSP8, respectively).

Statistical differences in GMT values among all the different evaluated times and proteins by group were assessed using a two-way analysis of variance (ANOVA) with Tukey’s multiple comparison test. Comparisons between CoronaVac-vaccinated individuals and convalescents subjects were assessed using the Kruskal-Wallis test with Dunn’s multiple comparisons test. The significance threshold was set at α = 0.05. GraphPad Prism 9.0 was used for data plotting, multiple logistic regression, and ROC curve statistical analyses.

## RESULTS

### Kinetics and extent of SARS-CoV-2-specific antibodies responses in convalescents, CoronaVac-vaccinated individuals, and breakthrough cases

To assess which are those antigens that contribute to the humoral response elicited after natural infection, vaccination, or infection after vaccination (breakthrough cases), first we performed dot blot analyses using a subset of non-structural (ORF1a, ORF3a, ORF8, NSP1, NSP8, NSP9, NSP10, NSP14,) and structural (E and M) recombinant viral proteins with serum pools from COVID-19 convalescent patients, CoronaVac-vaccinated individuals, and breakthrough cases. Naïve individuals were also included as controls. In these experiments, we detected some degree of differential immunoreactivity between the evaluated groups, mainly for the M and E structural proteins and the non-structural proteins ORF3a and NSP8 **(Supplementary Figure 1)**. We then evaluated the kinetics of the specific-IgG antibodies for these proteins and the structural N protein by ELISA in convalescent individuals, subjects vaccinated with two or three doses of CoronaVac, and breakthrough cases. Subject data and sampling times are detailed in Table 1.

As expected, CoronaVac-vaccinated individuals showed a humoral immune response that was polarized towards structural components within the virion **(Figure 1A-C).** Remarkably, N-specific IgG antibody titers were significantly elevated only four weeks after administering the third dose, compared to naïve controls, pre-immune samples, and sera obtained two weeks after the second dose. Conversely, M-specific antibodies titers showed a significant increase four weeks after the second dose, similar to those obtained after the booster dose **(Figure 1A).** There were no significant differences between vaccinated individuals and the negative controls for the NSP8 and ORF3a antigens **(Figure 1B).** The highest response in the vaccinated group was elicited against the N protein after the booster dose, which reached 87% seropositivity. In contrast, only around 50% of the individuals had N-specific antibodies after the second dose or M-specific antibodies after two or three doses of the vaccine. On the other hand, approximately 20% of the vaccinated individuals displayed a response against the NSP8 protein **(Figure 1C).**

**Figure 1.**
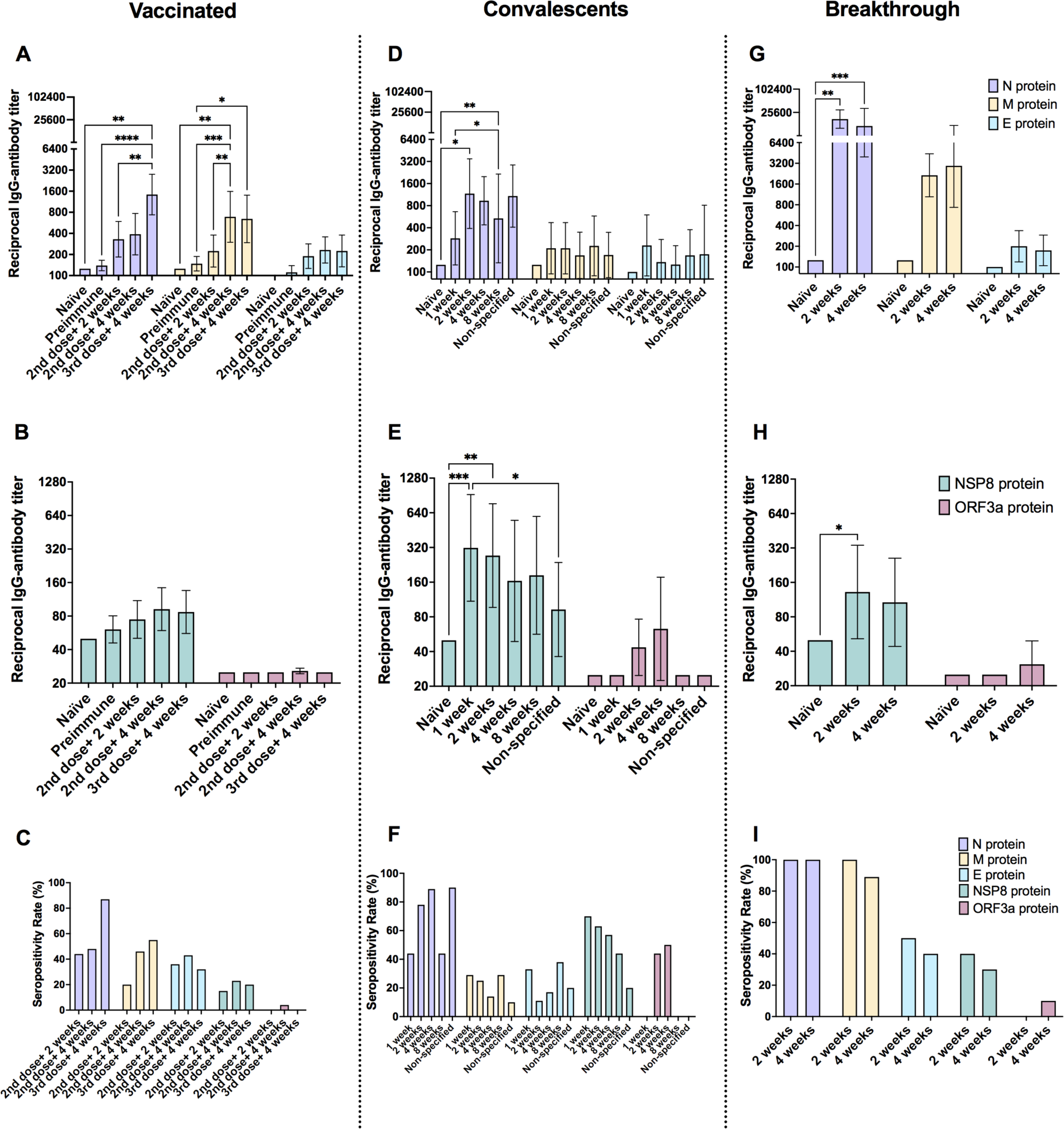
Kinetics of specific anti-IgG levels against multiple structural and non-structural SARS-CoV-2 proteins. A, B, and C. Serum samples from CoronaVac-vaccinated individuals were obtained at the moment of the first dose (pre-immune), two and four weeks after the second dose, and four weeks after a booster dose. **D, E, and F.** Convalescent individual serum sampling was performed at 1, 2, 4, and 8 weeks after recovering from SARS-CoV-2 infection. Nine convalescent individuals were evaluated without information about the time of sample collection (Unspecified). **G, H, and I.** Follow-up samples from CoronaVac breakthrough cases (right) were obtained two and four weeks after a positive PCR result. Serum from ten naïve individuals was added as controls. Reciprocal antibody titers elicited against structural SARS-CoV-2 proteins (N, M, and E) are shown in the upper panel. Reciprocal antibody titers elicited against non-structural SARS-CoV-2 proteins (NSP8 and ORF3a) are shown in the middle panel. The seropositivity rates were determined for each protein, and the respective time-points evaluated correspond to the lower panels. Bars show the Geometric Mean Titer (GMT), and the error bars indicate 95% CI. A two-way ANOVA test was used with Tukey’s multiple comparison post-test. *p < 0.05, **p < 0.01, ***p < 0.001, ****p < 0.0001.

Regarding the convalescent individuals, we detected significant differences in antibody titers between patients and naïve controls when assessing reactivity towards the N- structural protein and the NSP8-non-structural protein **(Figure 1D-F).** Sequential samples from some individuals showed that the antibody response against the N protein was significantly higher compared to naïve individuals starting two weeks after the onset of the symptoms and was maintained for up to eight weeks **(Figure 1D).** Moreover, we observed that the earliest antibody response in the convalescent group was elicited for the NSP8 protein after one and two weeks from the symptom’s onset **(Figure 1E).** The most immunogenic protein in this group was the N protein, with seropositivity rates ranging from 78 to 89% at two and four weeks after symptoms onset, respectively, followed by NSP8, which showed a seropositivity rate of 70 and 63% after one or two weeks, respectively **(Figure 1F).**

We next investigated the specific-IgG antibody responses produced in individuals with symptomatic SARS-CoV-2 infection who were previously vaccinated with two doses of CoronaVac (breakthrough cases) **(Figure 1G-I).** In this case, the N-specific IgG antibodies showed the highest levels at both times evaluated, two and four weeks after COVID-19 diagnosis, and showed 100% seropositivity rates. Although the antibody titers for the M protein did not reach significant levels as compared to naïve controls, they showed high seropositivity rates within 100 and 89% at two and four weeks after COVID-19 diagnosis, respectively **(Figures 1G and I).** To a lesser extent, we also found a significant difference at the antibody level between breakthrough cases and naïve controls for the NSP8 after two weeks, with a seropositivity rate of 40% (**Figures 1H** and I).

To harmonize the assessment of the humoral immune response after natural infection, vaccination, or in breakthrough cases, the WHO has recommended reporting the binding activity results in binding antibody units (BAU), using the first international standard (IS) for anti-SARS-CoV-2 immunoglobulin [29–31]. Antibody titers were obtained for the M, N, and NSP8 proteins as BAU (**Supplementary Figure 2 and Supplementary Table 1).** Antibody values for other viral proteins could not be reported as BAU, due to the low level of detection.

Overall, our results show that CoronaVac induces a broad humoral immune response, including high levels of N- and M-specific antibodies in an important percentage of vaccinated individuals, which was enhanced after natural infection in breakthrough cases. On the other hand, convalescent individuals who had not been vaccinated before infection developed a modest but statistically significant increase in antibody response against non-structural components of the virion.

### SARS-CoV-2-IgG responses among CoronaVac-vaccinated individuals, COVID-19 convalescent, and breakthrough subjects

To further investigate differences in the antibody responses between CoronaVac-vaccinated individuals and COVID-19 convalescent subjects, we analyzed the geometric mean titers (GMTs) elicited in each group, those vaccinated with two or three doses of CoronaVac, breakthrough cases after two doses of CoronaVac, and convalescent individuals without prior vaccination. Time-points evaluated were four weeks after the second or booster dose and four weeks after a positive PCR test for breakthrough cases. We also analyzed data from convalescent individuals that had sample collection with unspecified dates and samples from convalescent individuals whose samples were collected four weeks after symptom onset. A summary of the GMT values and seropositivity rate obtained at these time points are shown in **Table 2.**

**Table 2.** Seroconversion rates and geometric mean titers (GMTs) of antibodies against SARS-Cov-2 proteins

| **ANTIBODIES DETECTED** | **INDICATORS** | **2<sup>ND</sup> DOSE + 4 WEEKS** | **3<sup>RD</sup> DOSE + 4 WEEKS** | **BREAKTHROUGH + 4 WEEKS** | **CONVALESCENTS + 4 WEEKS** | **CONVALESCENTS UNSPECIFIED** |
| --- | --- | --- | --- | --- | --- | --- |
| **Anti-N IgG** | GMT | 389.58 | 1433.96 | 11313.71 | 933.03 | 1080.06 |
|  | 95% CI | 197.27-769.37 | 736.33 - 2792.54 | 3951.77 - 32390.55 | 438.27 - 1986.35 | 406.66 - 2868.58 |
|  | Seropositivity Rate (%) | 44 | 88 | 100 | 90 | 89 |
| **Anti-M IgG** | GMT | 688.50 | 643.33 | 2939.47 | 168.24 | 170.10 |
|  | 95% CI | 298.68 - 1587.12 | 295.06 - 1402.67 | 734.28 - 11767.36 | 81.33 - 348.02 | 83.59 - 346.12 |
|  | Seropositivity Rate (%) | 46 | 55 | 89 | 10 | 11 |
| **Anti-E IgG** | GMT | 232.03 | 224.49 | 174.11 | 125.99 | 174.11 |
|  | 95% CI | 151.12-356.25 | 133.11 - 378.60 | 104.33 - 290.56 | 69.57 - 228.18 | 37.34 - 811.82 |
|  | Seropositivity Rate (%) | 43 | 33 | 40 | 29 | 20 |
| **Anti-NSP8 IgG** | GMT | 92.31 | 87.06 | 107.18 | 164.07 | 92.59 |
|  | 95% CI | 59.26 - 143.80 | 55.80 - 135.82 | 44.08, 260.61 | 48.85, 551.01 | 36.18, 236.97 |
|  | Seropositivity Rate (%) | 27 | 24 | 30 | 57 | 22 |
| **Anti-ORF3a IgG** | GMT | 25.00 | 25.70 | 30.78 | 63.00 | 25.00 |
|  | 95% CI | 25.00 - 25.00 | 24.27 - 27.22 | 19.23 - 49.27 | 22.45 - 176.77 | 25.00 - 25.00 |
|  | Seropositivity Rate (%) | 0 | 5 | 10 | 50 | 0 |

Interestingly, the antibody responses were highest among breakthrough cases for the structural proteins N and M. As shown in **Figure 2,** significant differences of 11 and 18-fold were observed in terms of the magnitude of the anti-N and anti-M-specific IgG responses between breakthrough cases and convalescent individuals (GMTs: 11313.7 vs. 1000 and 2939.5 vs. 161.4, respectively). Importantly, convalescent individuals also had binding anti-M-specific IgG titers that were 4-fold lower than vaccinated individuals that received two or three vaccine doses, which reached similar GMTs of 688.5 and 643.3, respectively **(Figures 2A and B)**. We observed no significant difference in the overall levels of the anti-E-specific antibody responses in vaccinated individuals compared to convalescents, suggesting that infection or vaccination alone does not induce significant antibody responses against this protein **(Figure 2C).**

**Figure 2.**
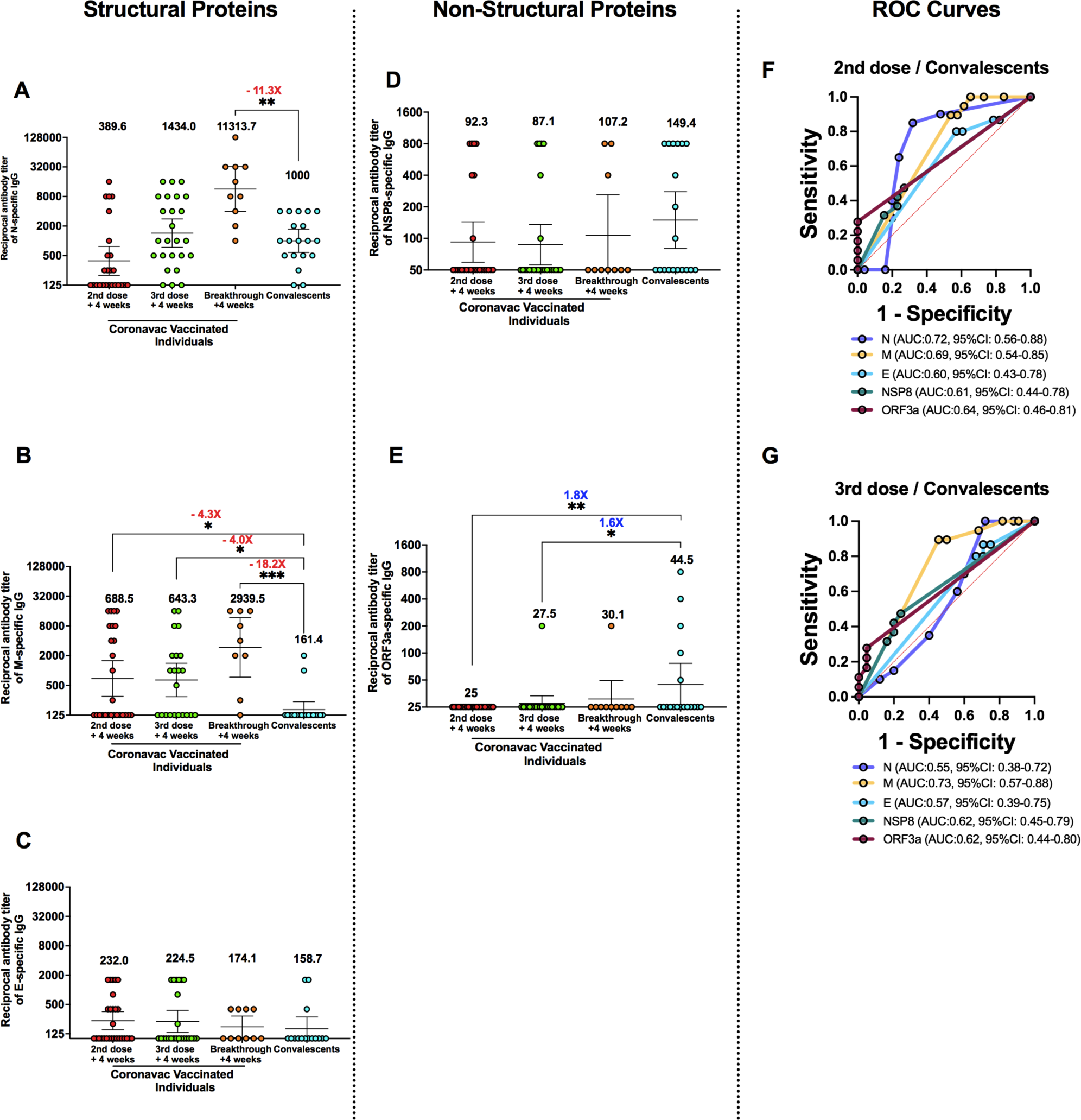
Comparison of antibody levels between convalescent subjects and CoronaVac-vaccinated individuals. Specific IgG antibodies titers in samples collected at four weeks after the second or booster dose in individuals belonging to the vaccinated group and four weeks after the diagnosis of SARS-CoV-2 infection by positive PCR result. Individuals from the breakthrough group were compared with those from the convalescent group (samples obtained four weeks after the recovery from SARS-CoV-2 infection and individuals for whom there is no data regarding when the samples were collected). **A, B, and C**. Reciprocal antibody titers against N, M, and E-specific IgG. **D and E.** Reciprocal IgG specific antibody titers against NSP-8 and ORF3a. The numbers above the groups show the Geometric Mean Titer (GMT), and the error bars indicate the 95% CI. The numbers in red indicate the fold decrease. The blue text indicates fold increases. A Kruskal-Wallis test was used with Dunn’s multiple comparison post-test, *p < 0.05, **p < 0.01, ***p < 0.001. **F and G.** ROC analyses of IgG responses to the N (violet), M (yellow), E (blue), NSP-8 (green), and ORF3a (pink) proteins to compare vaccinated individuals after the administration of two or three doses of CoronaVac and convalescent subjects, respectively. Ara under the curve (AUC) values are indicated in parenthesis.

Regarding the non-structural proteins evaluated, no differences were observed for the NSP8 protein, as some vaccinated individuals displayed high titers against this protein, and all groups reached GMTs of 92.3 and 87.1 after two or three vaccine doses, respectively. Likewise, breakthrough cases reached a GMT of 107.2 and the convalescent group a GMT of 149.4 **(Figure 2D).** These results suggest that vaccinated individuals may have had prior exposure to other coronaviruses that elicit cross-reactivity, displaying a nonspecific antibody immune response against this protein. However, this response was not boosted after natural exposure to SARS-CoV-2. By contrast, we detected significantly higher ORF3a-specific antibody titers in convalescent individuals than in CoronaVac-vaccinated subjects (GMT: 44.5 vs. 25 and 27.5 for two or three vaccine doses, respectively) **(Figure 2E)**.

Finally, we performed a receiver operating characteristic (ROC) analysis to identify which of the evaluated SARS-CoV-2 proteins may better differentiate CoronaVac-vaccinated from convalescent individuals. Using this approach, the best performance was obtained when testing structural proteins but depended on the number of doses administered to the subjects. On the one hand, high area under the curve (AUC) values were obtained for the N (0.72; 95%CI:0.56-0.88) and the M proteins (0.69; 95%CI:0.54-0.85) when comparing vaccinated individuals with CoronaVac-vaccinated individuals receiving only two doses and convalescents **(Figure 2F).** On the other hand, the N protein showed a low AUC value (0.55; 95%CI:0.38-0.72) when comparing vaccinated individuals receiving the third dose with convalescents, and in this case, on the contrary, the highest AUC value was achieved for the M protein (0.73; 95%CI:0.57-0.88) **(Figure 2G)**.

In summary, these results show that vaccinated individuals have a broader immune response and higher antibody titers against structural components of SARS-CoV-2 compared to individuals who underwent a natural infection without prior vaccination.

### Combination of N- and M-specific antibody responses for differentiating breakthrough cases from vaccinated-only, and convalescents individuals

As inactivated vaccines elicit a broad humoral immune response against various antigens, the serological diagnosis of previously vaccinated individuals and the evaluation of vaccine efficacy in a virus-exposed population is challenging. Next, we performed ROC analyses to identify SARS-CoV-2 proteins that may differentiate the antibody responses elicited in breakthrough cases from vaccinated-only individuals and convalescents subjects.

As shown in **Figure 3A,** when the humoral immune response elicited in breakthrough cases is compared with that elicited by individuals vaccinated with two doses of CoronaVac, we found AUC values of 0.92 (95%CI:0.83-1.0, N), 0.71 (95%CI:0.54-0.89, M), 0.55 (95%CI:0.35-0.75, E), 0.52 (95%CI:0.30-0.74, NSP8), and 0.55 (95%CI:0.33-0.77, ORF3a) for each protein, suggesting that M and N proteins could be useful for discriminating between both groups. We further compared individuals who received a booster dose (third dose) and found a similar tendency with the highest AUC values obtained for the N and M proteins, 0.83 (95%CI:0.68-0.97), and 0.76 (95%CI:0.58-0.94), respectively **(Figure 3B)**.

**Figure 3.**
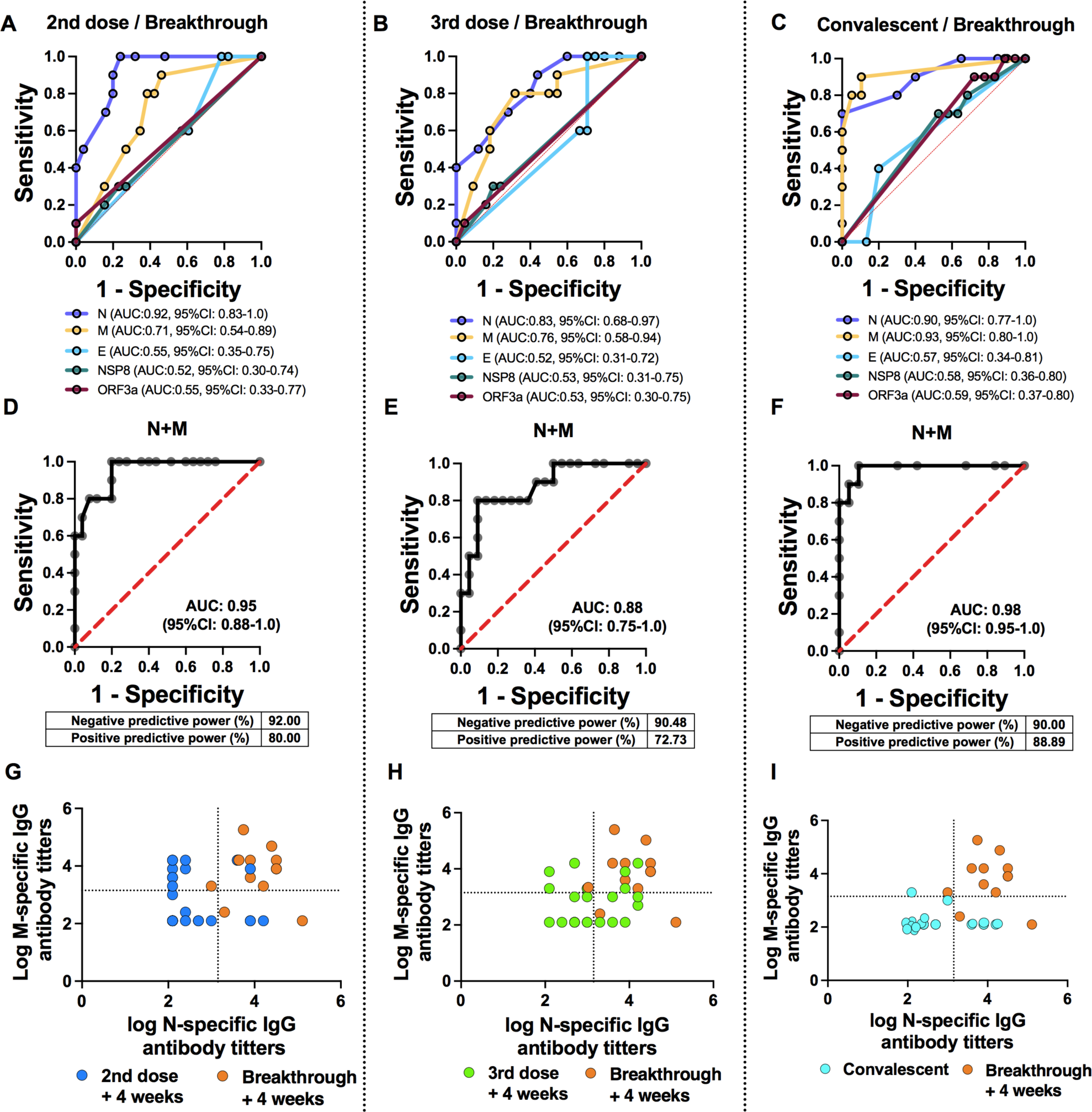
ROC analyses of IgG responses to structural and non-structural SARS-CoV- proteins for comparing CoronaVac breakthrough cases with vaccinated individuals and convalescent individuals. ROC curves were constructed to identify SARS-CoV-2 proteins that can differentiate breakthrough cases from **A.** individuals that received two doses of CoronaVac. **B.** individuals receiving three doses (two in the primary schedule, plus one booster) of CoronaVac **C.** Convalescent individuals. Sera collected four weeks after recovery from infection, vaccination, and breakthrough cases were analyzed. For the convalescent group, nine individuals with an unspecified time of sample collection were included. **D, E, and F.** AUC values, negative predictive power, and positive predictive power for the combinations of M and N antibodies titers to differentiate the breakthrough cases from **D.** individuals that received two doses of CoronaVac. **E.** individuals that received three doses of CoronaVac **F.** Convalescent individuals. **G, H and I**. Scatter plots distributions along with cutoff values for IgG responses based on optimal Youden values related to ROC curves: convalescent individuals (light blue), individuals vaccinated with two (dark blue), or three (green) doses, and breakthrough cases (orange).

Remarkably, these proteins are also able to differentiate infected subjects without prior vaccination (convalescents) from breakthrough cases (vaccinated, then infected), with AUC values up to 0.90 (95%CI:0.77-1.0), for the N protein and 0.93 (95%CI:0.81-1.0), for the M protein **(Figure 3C)**. In addition, we performed multiple logistic regression analyses to evaluate the capacity of the combined detection of N- and M-specific IgG antibodies to distinguish breakthrough cases from the other groups evaluated. As shown in **Figures 3D-F**, the combined data improved the AUC values obtained to 0.95 (95%CI:0.88-1.0), 0.88 (95%CI:0.75-1.0), and 0.98 (95%CI:0.95-1.0), respectively. Importantly, this approach showed high negative and positive predictive powers of 92% and 80% for differentiating breakthrough cases with vaccinated individuals that received two doses, and 90% and 73% when compared with subjects vaccinated with the booster dose, and lastly, 90% and 89% when comparing with convalescent individuals, suggesting this may be a valuable strategy for serological diagnosis and vaccine efficacy evaluation. Moreover, scatter plots performed with hypothetical cutoffs of IgG antibody titers showed that it is possible to cluster the majority of the breakthrough cases in the quadrant with the highest values for N and M antibody titers (Cutoff ≥1,500 M or N-specific IgG antibody titters), as compared with vaccinated-only individuals (two or three doses), or infected-only subjects (convalescents) **(Figures 3G-I).**

Overall, these results suggest that the combination of N- and M-specific IgG antibody levels could be a reliable biomarker for detecting vaccinated individuals who have had a mild or asymptomatic SARS-CoV-2 infection and potentially differentiate infected individuals from those with mixed immunity resulting from infection and vaccination with CoronaVac.

## DISCUSSION

In the context of the COVID-19 pandemic and mass vaccination campaigns being deployed around the world, a better understanding of the humoral immune response arising from either natural infection, vaccination, or infection after vaccination is highly relevant as a potential diagnostic tool that may help better determine epidemiological surveying and public health strategies. This may also help identify potential new correlates of protection against SARS-CoV-2 [32, 33]. For the vaccinated individuals, we analyzed the immune response elicited against a widely used inactivated SARS-CoV-2 vaccine (CoronaVac, Sinovac Biotech’s), given its significant deployment worldwide with billions of doses administered so far. Because this vaccine consists of whole inactivated SARS-CoV-2, it is composed of a diverse set of structural antigens that could be presented to the immune system, and thus it is somewhat expected to elicit a humoral immune response that may mirror natural infection [34].

Interestingly, antibodies raised against SARS-CoV-2 antigens were elicited at significantly different levels in individuals recovering from natural infection, after immunization with CoronaVac, and breakthrough cases. Furthermore, we found that antibody titers directed against the M-SARS-CoV-2 protein were significantly higher in the case of vaccinated individuals, as opposed to natural infection in unvaccinated individuals. Importantly, we observed high titers of anti-N antibodies in individuals boosted with an additional dose of inactivated SARS-CoV-2 vaccine, equivalent to those observed in convalescent individuals. Regarding breakthrough cases, we found a potent immune response for the two structural proteins N and M, suggesting that these humoral responses could be used to differentially detect immunity elicited after natural infection in this vaccinated group, as opposed to non-vaccinated infected individuals. Although we did not analyze potential correlations between the elicited antibody responses and protection against COVID-19, previous reports suggest that hybrid immunity, or additional antigen exposure after natural infection before or after vaccination, is associated with a lower risk of reinfection, as well as reduced COVID-19 hospitalization when compared to naturally acquired immunity after infection alone [35, 36]. Interestingly, high levels of IgG against the N protein, at the time of hospital admission, has been correlated with worsen COVID-19 clinical courses [37], which could be attributed to increased IL-6 production mediated by N-specific IgG antibodies observed after COVID-19 cytokine storm [38]. On the contrary, high levels of IgG against the N protein, detected after COVID-19 recovery, have been associated with a long-term protective effect against re-infections [39]. The effect of other antibodies on the outcome of COVID-19 has not been explored to date.

On the other hand, antibody titers against the ORF3a protein seemed to be elicited almost exclusively in response to natural infection in non-vaccinated individuals (convalescents) [40, 41]. Therefore, ORF3a-SARS-CoV-2 may represent a biomarker to differentiate natural SARS-CoV-2 infection from vaccination. Noteworthy, only a fraction (50%) of the infected individuals responded to this antigen, which should be considered for further analyses. Furthermore, NSP8-specific antibodies were elicited in a great percentage of convalescent individuals at early time points (70% during the first week after symptom onset). Although some individuals in the vaccinated group responded to this antigen, it is unclear whether they were previously exposed to the virus before vaccination or whether it corresponds to potential antibody cross-reactivity against other coronaviruses. However, these findings need to be corroborated in future larger-scale studies to confirm the diagnostic value of this protein and its potential use as a biomarker for SARS-CoV-2 infection.

Importantly, as discussed in the preceding paragraph, our results show that immunization with CoronaVac, namely three doses, robustly elicits antibodies against the N and M proteins of SARS-CoV-2 in human volunteers, which to our knowledge has not been reported before, and which is consistent with the fact that inactivated SARS-CoV-2 vaccines present a plethora of antigens to the immune system that elicits antibodies against multiple viral proteins. Notably, as viral variants with multiple mutations in the S protein continue to emerge, it is of considerable importance the induction of polyclonal responses elicited by inactivated vaccines that could help to avoid the viral escape [10]. These data complement our previous findings that reported high titers of anti-S-RBD antibodies and neutralizing antibodies against SARS-CoV-2 in phase III clinical trials on adult and pediatric volunteers [9].

On the other hand, our results complement findings previously reported for the SARS-CoV-2 inactivated virus vaccine BBIBP-CorV, where the authors showed that the N protein, NSP7, and S2-78 peptide may be applied independently or in combination for effective discrimination between individuals vaccinated with two doses and convalescents subjects. However, individuals vaccinated with three doses or breakthrough cases were not included, and M-specific IgG response was not investigated [42]. In another study, the combined use of ORF3b and ORF8 showed to be accurate serological markers for detection of early and late SARS-CoV-2 infection, but further studies evaluating their potential to differentiate convalescent and vaccinated individuals at different time points after vaccination and/or with different vaccines are missing [43]. Moreover, the specific properties of the antibodies elicited after vaccination or natural infection, other than neutralization capacity, should be considered in future studies to accurately identify a correlate of protection against COVID-19 [36, 37].

Overall, our results indicate that CoronaVac is immunogenic and that the host can produce various antibodies against structural proteins of SARS-CoV-2. This could allow for a potential differential diagnosis based on humoral immune components present in the sera among subjects undergoing natural infection, vaccinated, or breakthrough cases. Moreover, our data show that immunization with an inactivated SARS-CoV-2 vaccine induces significantly higher levels of antibodies against the structural protein M-SARS-CoV-2 compared to natural infection in unvaccinated subjects (convalescent subjects). Interestingly, natural infection with SARS-CoV-2 elicits significantly higher titers of these antibodies when the individuals were previously immunized (breakthrough cases) compared to convalescent subjects. Additionally, we identified an antigen, ORF3a-SARS-CoV-2, against which antibodies were produced in convalescent subjects at significantly higher titers than in CoronaVac-immunized individuals.

Finally, it will be interesting to confirm the findings reported herein using high throughput analyses that consider a significant number of samples accompanied with the corresponding demographic information and details of the timing of the infection and severity of the reported symptoms for non-vaccinated convalescent individuals and breakthrough cases. Similarly, knowing the SARS-CoV-2 variant infecting the individuals could shed light onto potential differences regarding the immunogenicity of these viruses.

Taken together, our study reveals different SARS-CoV-2 antigens that could be used as differential biomarkers, alone or combined among them, for distinguishing between natural infection-, vaccination-elicited immune memory, or infection after vaccination. Here, we report that the structural M and N proteins are produced in a high percentage of vaccinated individuals along with high titers, which are further enhanced after a natural infection. These data could be helpful not only for serodiagnosis but also for evaluating vaccine efficacy through blood sample determination in the actual context in which SARS-CoV-2 infection is highly prevalent worldwide.

## Data Availability

All data produced in the present work are contained in the manuscript.

## ACKNOWLEDGMENTS

Special thanks to Rami Scharf, Miren Iturriza-Gomara and Jorge Flores from PATH for their support on experimental design and discussion. We also thank the Vice Presidency of Research (VRI), the Direction of Technology Transfer and Development (DTD), the Legal Affairs Department (DAJ) of the Pontificia Universidad Católica de Chile. We are also grateful to the Administrative Directions of the School of Biological Sciences (FCB) and the School of Medicine (FMed) of the Pontificia Universidad Católica de Chile for their administrative support.

## SUPPLEMENTARY TABLES AND FIGURES

**Supplementary Table 1.** Geometric mean units (GMU) of antibodies against SARS-CoV-2 proteins

| ANTIBODIES DETECTED | INDICATORS | 2<sup>ND</sup> DOSE<br>+ 4 WEEKS | 3<sup>RD</sup> DOSE<br>+ 4 WEEKS | BREAKTHROUGH<br>+4 WEEKS | CONVALESCENTS<br>+ 4 WEEKS | CONVALESCENTS<br>UNSPECIFIED<br>INFECTION TIME |
| --- | --- | --- | --- | --- | --- | --- |
| Anti-N IgG | GMU | 79.1 | 197.2 | 1224.0 | 127.0 | 116.7 |
|  | 95% CI | 41.75- 150.2 | 96.7 - 402.1 | 495.4 - 3026 | 67.84 - 237.7 | 50.74 - 268.2 |
| Anti- M IgG | GMU | 246.1 | 163.9 | 586.0 | 51.26 | 31.9 |
|  | 95% CI | 136.3 - 444.2 | 94.75 -283.4 | 234.1 - 1467 | 27.53 - 95.46 | 15.92 - 63.89 |
| Anti-NSP8<br>IgG | GMU | 15.5 | 14.68 | 20.5 | 20.66 | 23.62 |
|  | 95% CI | 13.2- 18.31 | 13.0- 16.57 | 10.4- 40.4 | 12.5- 34.11 | 10.0- 55.59 |

**Figure S1.**
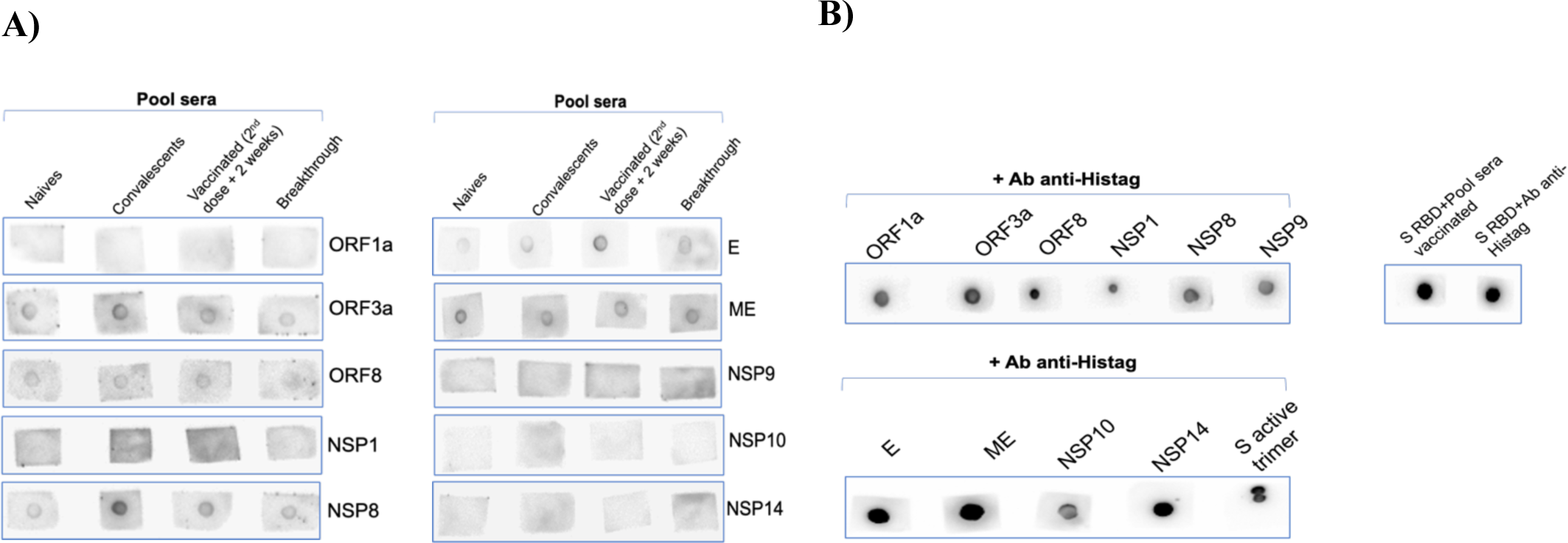
Anti-SARS-COV-2 antibodies were detected by Dot Blot. Five hundred nanograms (500 ng) of SARS-CoV-2 recombinant proteins (ORF1a, ORF3a, ORF8, NSP1, NSP8, NSP9, NSP10, NSP14, E (envelop), ME (membrane)) were immobilized in nitrocellulose membranes using 2 uL of proteins prepared in 8M Urea denaturant buffer. The membranes were blocked with 10% BSA and incubated overnight at 4°C with a sera pool obtained from control naïve individuals, convalescents (PCR (+) + 4w), CoronaVac-vaccinated with 2nd dose + 2 weeks, and breakthrough (PCR (+) + 2 weeks) subjects (dilution 1/250). As positive controls, all proteins (500 ng) were incubated with an anti-HisTag antibody conjugated with biotin. After incubation with sera, the membranes were treated with anti-human IgG-HRP. Finally, the membranes were incubated with an enhanced chemiluminescence Western blot detection system (Femto, ECL).

**Figure S2.**
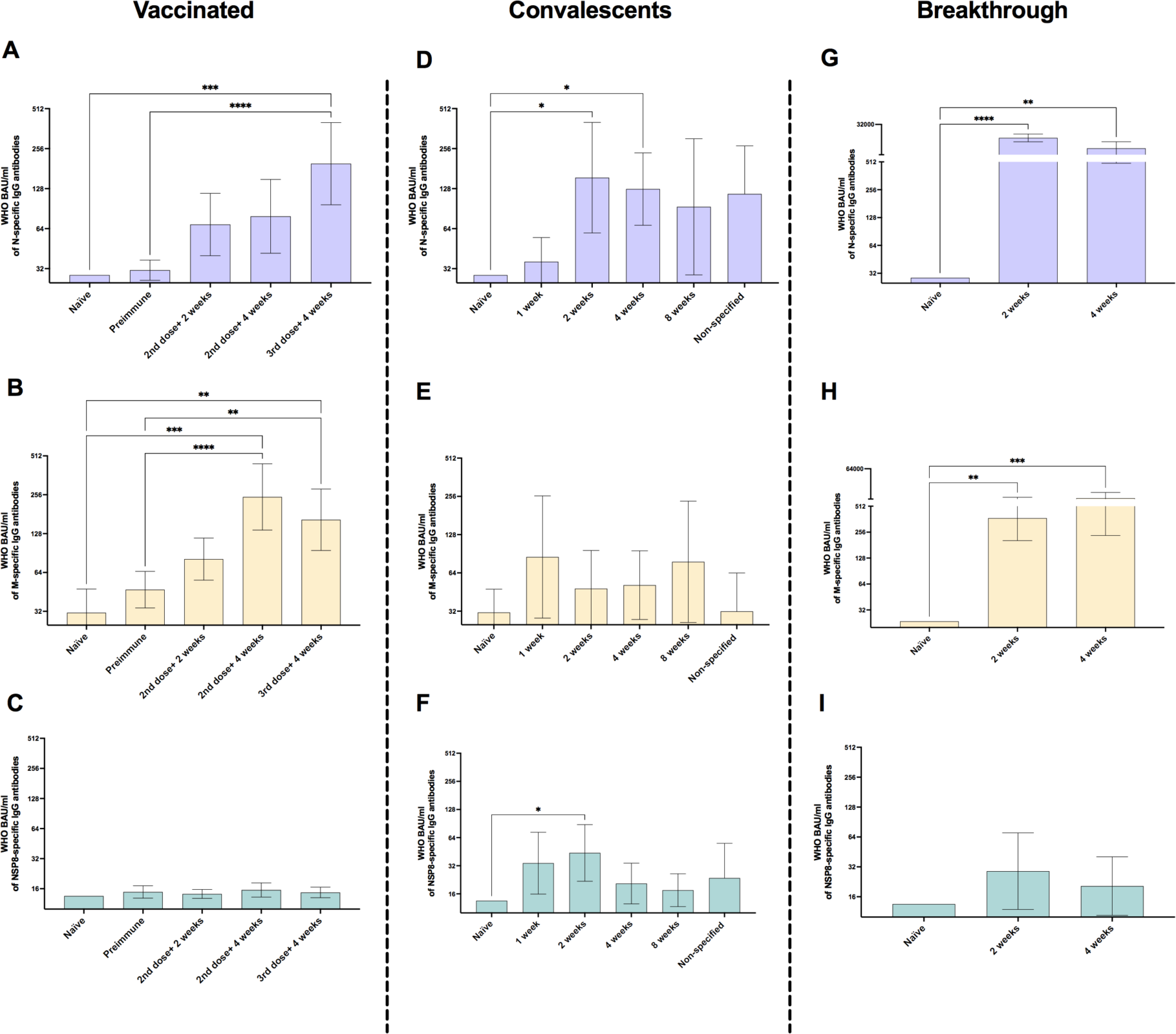
IgG level kinetics against structural and non-structural SARS-CoV-2 proteins expressed as WHO-binding antibody units (BAU). Serum samples from CoronaVac-vaccinated individuals were obtained at the first dose (pre-immune), two and four weeks after the second dose, and four weeks after a booster dose for evaluating **A.** N-specific IgG antibodies, **B.** M-specific IgG antibodies, **C.** NSP8-specific IgG antibodies. Serum sampling of convalescent individuals was performed at 1, 2, 4, and 8 weeks after recovery from SARS-CoV-2 infection. Nine convalescent individuals without information regarding the time of sample collection (Unspecified) were included for evaluating, **D.** N-specific IgG antibodies, **E.** M-specific IgG antibodies, **F.** NSP8-specific IgG antibodies. Follow-up samples from breakthrough cases were obtained two and four weeks after a positive PCR result for evaluating **G.** N-specific IgG antibodies, **H.** M-specific IgG antibodies, and **I.** NSP8-specific IgG antibodies. Sera from ten naïve individuals were added as controls. Bars show the Geometric Mean Units (GMU), and the error bars indicate 95% CI. A Kruskal-Wallis test was used with Dunn’s multiple comparison post-test, *p < 0.05, **p < 0.01, ***p < 0.001, ****p < 0.0001.

## REFERENCES

1. World Health Organization Global Excess Deaths Associated with COVID-19, January 2020 - December 2021.

2. Richards, F.; Kodjamanova, P.; Chen, X.; Li, N.; Atanasov, P.; Bennetts, L.; Patterson, B.J.; Yektashenas, B.; Mesa-Frias, M.; Tronczynski, K.;, et al. Economic Burden of COVID-19: A Systematic Review. ClinicoEconomics and Outcomes Research 2022, *Volume* 14, 293–307, doi:10.2147/CEOR.S338225.

3. Krammer, F. A Correlate of Protection for SARS-CoV-2 Vaccines Is Urgently Needed. Nature Medicine 2021, 27, 1147–1148, doi:10.1038/s41591-021-01432-4.

4. Nagy, A.; Alhatlani, B. An Overview of Current COVID-19 Vaccine Platforms. Computational and Structural Biotechnology Journal 2021, 19, 2508–2517, doi:10.1016/j.csbj.2021.04.061.

5. Al-Jighefee, H.T.; Najjar, H.; Ahmed, M.N.; Qush, A.; Awwad, S.; Kamareddine, L. COVID-19 Vaccine Platforms: Challenges and Safety Contemplations. Vaccines (Basel) 2021, 9, 1196, doi:10.3390/vaccines9101196.

6. Ranzani, O.T.; Hitchings, M.D.T.; Dorion, M.; D’Agostini, T.L.; de Paula, R.C.; de Paula, O.F.P.; Villela, E.F. de M.; Torres, M.S.S.; de Oliveira, S.B.; Schulz, W.;, et al. Effectiveness of the CoronaVac Vaccine in Older Adults during a Gamma Variant Associated Epidemic of Covid-19 in Brazil: Test Negative Case-Control Study. BMJ 2021, n2015, doi:10.1136/bmj.n2015.

7. Zhang, Y.; Zeng, G.; Pan, H.; Li, C.; Hu, Y.; Chu, K.; Han, W.; Chen, Z.; Tang, R.; Yin, W.;, et al. Safety, Tolerability, and Immunogenicity of an Inactivated SARS-CoV-2 Vaccine in Healthy Adults Aged 18–59 Years: A Randomised, Double-Blind, Placebo-Controlled, Phase 1/2 Clinical Trial. The Lancet Infectious Diseases 2021, 21, 181–192, doi:10.1016/S1473-3099(20)30843-4.

8. Wu, Z.; Hu, Y.; Xu, M.; Chen, Z.; Yang, W.; Jiang, Z.; Li, M.; Jin, H.; Cui, G.; Chen, P.;, et al. Safety, Tolerability, and Immunogenicity of an Inactivated SARS-CoV-2 Vaccine (CoronaVac) in Healthy Adults Aged 60 Years and Older: A Randomised, Double-Blind, Placebo-Controlled, Phase 1/2 Clinical Trial. The Lancet Infectious Diseases 2021, 21, 803–812, doi:10.1016/S1473-3099(20)30987-7.

9. Bueno, S.M.; Abarca, K.; González, P.A.; Gálvez, N.M.S.; Soto, J.A.; Duarte, L.F.; Schultz, B.M.; Pacheco, G.A.; González, L.A.; Vázquez, Y.;, et al. Safety and Immunogenicity of an Inactivated Severe Acute Respiratory Syndrome Coronavirus 2 Vaccine in a Subgroup of Healthy Adults in Chile. Clinical Infectious Diseases 2021, doi:10.1093/cid/ciab823.

10. Melo-González, F.; Soto, J.A.; González, L.A.; Fernández, J.; Duarte, L.F.; Schultz, B.M.; Gálvez, N.M.S.; Pacheco, G.A.; Ríos, M.; Vázquez, Y.;, et al. Recognition of Variants of Concern by Antibodies and T Cells Induced by a SARS-CoV-2 Inactivated Vaccine. Frontiers in Immunology 2021, 12, doi:10.3389/fimmu.2021.747830.

11. Jara, A.; Undurraga, E.A.; González, C.; Paredes, F.; Fontecilla, T.; Jara, G.; Pizarro, A.; Acevedo, J.; Leo, K.; Leon, F.;, et al. Effectiveness of an Inactivated SARS-CoV-2 Vaccine in Chile. New England Journal of Medicine 2021, 385, 875–884, doi:10.1056/NEJMoa2107715.

12. Ranzani, O.T.; Hitchings, M.D.T.; Dorion, M.; D’Agostini, T.L.; de Paula, R.C.; de Paula, O.F.P.; Villela, E.F. de M.; Torres, M.S.S.; de Oliveira, S.B.; Schulz, W.;, et al. Effectiveness of the CoronaVac Vaccine in Older Adults during a Gamma Variant Associated Epidemic of Covid-19 in Brazil: Test Negative Case-Control Study. BMJ 2021, n2015, doi:10.1136/bmj.n2015.

13. Duarte, L.F.; Gálvez, N.M.S.; Iturriaga, C.; Melo-González, F.; Soto, J.A.; Schultz, B.M.; Urzúa, M.; González, L.A.; Vázquez, Y.; Ríos, M.;, et al. Immune Profile and Clinical Outcome of Breakthrough Cases After Vaccination With an Inactivated SARS-CoV-2 Vaccine. Frontiers in Immunology 2021, 12, doi:10.3389/fimmu.2021.742914.

14. Tanriover, M.D.; Doğanay, H.L.; Akova, M.; Güner, H.R.; Azap, A.; Akhan, S.; Köse, Ş.; Erdinç, F.Ş.; Akalın, E.H.; Tabak, Ö.F.;, et al. Efficacy and Safety of an Inactivated Whole-Virion SARS-CoV-2 Vaccine (CoronaVac): Interim Results of a Double-Blind, Randomised, Placebo-Controlled, Phase 3 Trial in Turkey. The Lancet 2021, 398, 213–222, doi:10.1016/S0140-6736(21)01429-X.

15. Alencar, C.H.; Cavalcanti, L.P. de G.; Almeida, M.M. de; Barbosa, P.P.L.; Cavalcante, K.K. de S.; Melo, D.N. de; de Brito Alves, B.C.F.; Heukelbach, J. High Effectiveness of SARS-CoV-2 Vaccines in Reducing COVID-19-Related Deaths in over 75-Year-Olds, Ceará State, Brazil. Tropical Medicine and Infectious Disease 2021, 6, 129, doi:10.3390/tropicalmed6030129.

16. Fiori, M.; Bello, G.; Wschebor, N.; Lecumberry, F.; Ferragut, A.; Mordecki, E. Decoupling between SARS-CoV-2 Transmissibility and Population Mobility Associated with Increasing Immunity from Vaccination and Infection in South America. Scientific Reports 2022, 12, 6874, doi:10.1038/s41598-022-10896-4.

17. Costa Clemens, S.A.; Weckx, L.; Clemens, R.; Almeida Mendes, A.V.; Ramos Souza, A.; Silveira, M.B. v; da Guarda, S.N.F.; de Nobrega, M.M.; de Moraes Pinto, M.I.; Gonzalez, I.G.S.;, et al. Heterologous versus Homologous COVID- 19 Booster Vaccination in Previous Recipients of Two Doses of CoronaVac COVID-19 Vaccine in Brazil (RHH-001): A Phase 4, Non-Inferiority, Single Blind, Randomised Study. The Lancet 2022, 399, 521–529, doi:10.1016/S0140-6736(22)00094-0.

18. Farias, J.P.; da Silva, P. de S.; Fogaça, M.M.C.; Santana, I.V.R.; Luiz, W.B.; Birbrair, A.; Amorim, J.H. The COVID-19 Humoral Immunological Status Induced by CoronaVac and AstraZeneca Vaccines Significantly Benefits from a Booster Shot with the Pfizer Vaccine. Journal of Virology 2022, doi:10.1128/jvi.00177-22.

19. Jara, A.; Undurraga, E.A.; Zubizarreta, J.R.; González, C.; Pizarro, A.; Acevedo, J.; Leo, K.; Paredes, F.; Bralic, T.; Vergara, V.;, et al. Effectiveness of Homologous and Heterologous Booster Doses for an Inactivated SARS-CoV-2 Vaccine: A Large-Scale Prospective Cohort Study. The Lancet Global Health 2022, 10, e798–e806, doi:10.1016/S2214-109X(22)00112-7.

20. Tabilo Valenzuela, P.B.; Flores Balter, G.; Saint-Pierre Contreras, G.; Conei Valencia, D.; Moreno Calderón, C.; Bohle Venegas, C.; Guajardo Rivera, M.; Silva Ojeda, F.; Vial Covarrubias, M.J. Cellular Immune Response in Patients Immunized with Three Vaccine Doses of Different Vaccination Schemes Authorized by the Chilean Ministry of Health in January 2022. Life 2022, 12, 534, doi:10.3390/life12040534.

21. Redondo, N.; Zaldívar-López, S.; Garrido, J.J.; Montoya, M. SARS-CoV-2 Accessory Proteins in Viral Pathogenesis: Knowns and Unknowns. Frontiers in Immunology 2021, 12, doi:10.3389/fimmu.2021.708264.

22. Arya, R.; Kumari, S.; Pandey, B.; Mistry, H.; Bihani, S.C.; Das, A.; Prashar, V.; Gupta, G.D.; Panicker, L.; Kumar, M. Structural Insights into SARS-CoV-2 Proteins. Journal of Molecular Biology 2021, 433, 166725, doi:10.1016/j.jmb.2020.11.024.

23. Xia, X. Domains and Functions of Spike Protein in SARS-Cov-2 in the Context of Vaccine Design. Viruses 2021, 13, 109, doi:10.3390/v13010109.

24. Ke, Z.; Oton, J.; Qu, K.; Cortese, M.; Zila, V.; McKeane, L.; Nakane, T.; Zivanov, J.; Neufeldt, C.J.; Cerikan, B.;, et al. Structures and Distributions of SARS-CoV-2 Spike Proteins on Intact Virions. Nature 2020, 588, 498–502, doi:10.1038/s41586-020-2665-2.

25. Romero Ramírez, D.S.; Lara Pérez, M.M.; Carretero Pérez, M.; Suárez Hernández, M.I.; Martín Pulido, S.; Pera Villacampa, L.; Fernández Vilar, A.M.; Rivero Falero, M.; González Carretero, P.; Reyes Millán, B.;, et al. SARS-CoV-2 Antibodies in Breast Milk After Vaccination. Pediatrics 2021, 148, doi:10.1542/peds.2021-052286.

26. Barnes, C.O.; Jette, C.A.; Abernathy, M.E.; Dam, K.-M.A.; Esswein, S.R.; Gristick, H.B.; Malyutin, A.G.; Sharaf, N.G.; Huey-Tubman, K.E.; Lee, Y.E.;, et al. SARS-CoV-2 Neutralizing Antibody Structures Inform Therapeutic Strategies. Nature 2020, 588, 682–687, doi:10.1038/s41586-020-2852-1.

27. Tea, F.; Ospina Stella, A.; Aggarwal, A.; Ross Darley, D.; Pilli, D.; Vitale, D.; Merheb, V.; Lee, F.X.Z.; Cunningham, P.; Walker, G.J.;, et al. SARS-CoV-2 Neutralizing Antibodies: Longevity, Breadth, and Evasion by Emerging Viral Variants. PLOS Medicine 2021, 18, e1003656, doi:10.1371/journal.pmed.1003656.

28. World Health Organization WHO/BS.2020.2403 Establishment of the WHO International Standard and Reference Panel for Anti-SARS-CoV-2 Antibody.

29. Knezevic, I.; Mattiuzzo, G.; Page, M.; Minor, P.; Griffiths, E.; Nuebling, M.; Moorthy, V. WHO International Standard for Evaluation of the Antibody Response to COVID-19 Vaccines: Call for Urgent Action by the Scientific Community. The Lancet Microbe 2022, 3, e235–e240, doi:10.1016/S2666-5247(21)00266-4.

30. Infantino, M.; Pieri, M.; Nuccetelli, M.; Grossi, V.; Lari, B.; Tomassetti, F.; Calugi, G.; Pancani, S.; Benucci, M.; Casprini, P.;, et al. The WHO International Standard for COVID-19 Serological Tests: Towards Harmonization of Anti-Spike Assays. International Immunopharmacology 2021, 100, 108095, doi:10.1016/j.intimp.2021.108095.

31. Kristiansen, P.A.; Page, M.; Bernasconi, V.; Mattiuzzo, G.; Dull, P.; Makar, K.; Plotkin, S.; Knezevic, I. WHO International Standard for Anti-SARS-CoV-2 Immunoglobulin. The Lancet 2021, 397, 1347–1348, doi:10.1016/S0140-6736(21)00527-4.

32. Earle, K.A.; Ambrosino, D.M.; Fiore-Gartland, A.; Goldblatt, D.; Gilbert, P.B.; Siber, G.R.; Dull, P.; Plotkin, S.A. Evidence for Antibody as a Protective Correlate for COVID-19 Vaccines. Vaccine 2021, 39, 4423–4428, doi:10.1016/j.vaccine.2021.05.063.

33. Khoury, D.S.; Cromer, D.; Reynaldi, A.; Schlub, T.E.; Wheatley, A.K.; Juno, J.A.; Subbarao, K.; Kent, S.J.; Triccas, J.A.; Davenport, M.P. Neutralizing Antibody Levels Are Highly Predictive of Immune Protection from Symptomatic SARS-CoV-2 Infection. Nature Medicine 2021, 27, 1205–1211, doi:10.1038/s41591-021-01377-8.

34. Sadarangani, M.; Marchant, A.; Kollmann, T.R. Immunological Mechanisms of Vaccine-Induced Protection against COVID-19 in Humans. Nature Reviews Immunology 2021, 21, 475–484, doi:10.1038/s41577-021-00578-z.

35. Bates, T.A.; McBride, S.K.; Leier, H.C.; Guzman, G.; Lyski, Z.L.; Schoen, D.; Winders, B.; Lee, J.-Y.; Lee, D.X.; Messer, W.B.;, et al. Vaccination before or after SARS-CoV-2 Infection Leads to Robust Humoral Response and Antibodies That Effectively Neutralize Variants. Science Immunology 2022, 7, doi:10.1126/sciimmunol.abn8014.

36. Nordström, P.; Ballin, M.; Nordström, A. Risk of SARS-CoV-2 Reinfection and COVID-19 Hospitalisation in Individuals with Natural and Hybrid Immunity: A Retrospective, Total Population Cohort Study in Sweden. The Lancet Infectious Diseases 2022, 22, 781–790, doi:10.1016/S1473-3099(22)00143-8.

37. Batra, M.; Tian, R.; Zhang, C.; Clarence, E.; Sacher, C.S.; Miranda, J.N.; de La Fuente, J.R.O.; Mathew, M.; Green, D.; Patel, S.;, et al. Role of IgG against N-Protein of SARS-CoV2 in COVID19 Clinical Outcomes. Scientific Reports 2021, 11, 3455, doi:10.1038/s41598-021-83108-0.

38. Nakayama, E.E.; Kubota-Koketsu, R.; Sasaki, T.; Suzuki, K.; Uno, K.; Shimizu, J.; Okamoto, T.; Matsumoto, H.; Matsuura, H.; Hashimoto, S.;, et al. Anti-Nucleocapsid Antibodies Enhance the Production of IL-6 Induced by SARS-CoV-2 N Protein. Scientific Reports 2022, 12, 8108, doi:10.1038/s41598-022-12252-y.

39. Kohler, P.; Güsewell, S.; Seneghini, M.; Egger, T.; Leal, O.; Brucher, A.; Lemmenmeier, E.; Möller, J.C.; Rieder, P.; Ruetti, M.;, et al. Impact of Baseline SARS-CoV-2 Antibody Status on Syndromic Surveillance and the Risk of Subsequent COVID-19—a Prospective Multicenter Cohort Study. BMC Medicine 2021, 19, 270, doi:10.1186/s12916-021-02144-9.

40. Dugan, H.L.; Stamper, C.T.; Li, L.; Changrob, S.; Asby, N.W.; Halfmann, P.J.; Zheng, N.-Y.; Huang, M.; Shaw, D.G.; Cobb, M.S.;, et al. Profiling B Cell Immunodominance after SARS-CoV-2 Infection Reveals Antibody Evolution to Non-Neutralizing Viral Targets. Immunity 2021, 54, 1290–1303.e7, doi:10.1016/j.immuni.2021.05.001.

41. Cheng, L.; Zhang, X.; Chen, Y.; Wang, D.; Zhang, D.; Yan, S.; Wang, H.; Xiao, M.; Liang, T.; Li, H.;, et al. Dynamic Landscape Mapping of Humoral Immunity to SARS-CoV-2 Identifies Non-Structural Protein Antibodies Associated with the Survival of Critical COVID-19 Patients. Signal Transduction and Targeted Therapy 2021, 6, 304, doi:10.1038/s41392-021-00718-w.

42. Ma, M.-L.; Shi, D.-W.; Li, Y.; Hong, W.; Lai, D.-Y.; Xue, J.-B.; Jiang, H.-W.; Zhang, H.-N.; Qi, H.; Meng, Q.-F.;, et al. Systematic Profiling of SARS-CoV- 2-Specific IgG Responses Elicited by an Inactivated Virus Vaccine Identifies Peptides and Proteins for Predicting Vaccination Efficacy. Cell Discovery 2021, 7, 67, doi:10.1038/s41421-021-00309-7.

43. Hachim, A.; Kavian, N.; Cohen, C.A.; Chin, A.W.H.; Chu, D.K.W.; Mok, C.K.P.; Tsang, O.T.Y.; Yeung, Y.C.; Perera, R.A.P.M.; Poon, L.L.M.;, et al. ORF8 and ORF3b Antibodies Are Accurate Serological Markers of Early and Late SARS-CoV-2 Infection. Nature Immunology 2020, 21, 1293–1301, doi:10.1038/s41590-020-0773-7.

